# Metagenomic profiling of long-read sequencing for clinical diagnosis of ocular inflammation

**DOI:** 10.1101/2023.01.20.23284817

**Authors:** Yoshito Koyanagi, Ai Fujita Sajiki, Hiroaki Ushida, Kenichi Kawano, Kosuke Fujita, Daishi Okuda, Mitsuki Kawabe, Kazuhisa Yamada, Ayana Suzumura, Shu Kachi, Hiroki Kaneko, Hiroyuki Komatsu, Yoshihiko Usui, Hiroshi Goto, Koji M. Nishiguchi

## Abstract

**Objective:** To establish a metagenomic profiling method using long-read sequencing for clinical diagnosis of ocular inflammation and detect the etiologic virus of herpetic uveitis.

**Design:** A retrospective, cross-sectional study.

**Participants:** The participants were 44 uveitis patients with a suspected infectious etiology and 22 controls with cataract.

**Methods:** The anterior aqueous humor (10-20 µl) was subjected to DNA purification, followed by whole genome amplification. The Nanopore MinION™ using the Flongle Flow Cell was used to perform rapid long-read sequencing and the phylogenetic composition of the microorganisms in the specimen was evaluated.

**Main Outcomes and Measures:** The detection of the DNA sequence reads of the etiologic virus of herpetic uveitis in the generated FASTQ files from nanopore sequencing and the evaluation of the limits of detection (LOD) of metagenomic analysis compared to multiplex polymerase chain reaction (mPCR) testing for etiologic virus detection of herpetic uveitis.

**Results:** The detection rate of nanopore metagenomic analysis was approximately 59.0% as a result of validation against 22 mPCR-positive cases. The LOD was between 10^3.6^ and 10^6^ copies of virus DNA. The undetectable cases tended to have significantly lower copy numbers by mPCR, suggesting the lower metagenomic analysis sensitivity compared to mPCR. The nine pathogenic microorganisms evaluated by mPCR were also not detected by nanopore in all mPCR-negative cases and controls. The minimum time to obtain analysis results using this method was approximately 190 minutes.

**Conclusions and Relevance:** Our established sequencing protocol from the anterior aqueous humor detected the DNA fragments of etiologic viruses in patients with herpes virus uveitis. Conversely, nanopore metagenomic results contained considerable noise and were found to be less sensitive compared to the conventional mPCR tests for ocular infections.

## Introduction

In the field of ophthalmology, multiplex polymerase chain reaction (mPCR) testing is commonly used to detect the major pathogenic microorganisms from intraocular fluid (approximately 20 μl) and corneal abrasion specimens. The concordance rate with conventional quantitative PCR (qPCR) is high (98.8%–100%), contributing to the improvement of the ocular infectious diseases diagnosis rate, especially herpesvirus infections.^1-3^ However, even with the widespread use of this method, infectious uveitis accounts for only 16% of all uveitis cases and the cause of inflammation remains unidentified and undiagnosed in 37% of all cases in Japan.^4^ Improving the ocular infection diagnosis rate is a topic of great clinical importance because most infections can be treated with antibiotics and antivirals and the treatment strategy is fundamentally different from that of noninfectious diseases, which primarily suppresses inflammation.

Recently, metagenomic analysis has become widely used in the field of bacteriology. It is defined as the direct purification and comprehensive DNA and RNA sequencing of microbial communities in specimens without the process of culturing them to obtain information on the microorganisms present. Conventional mPCR is a method to specifically detect the genome of a known pathogen, whereas metagenomic analysis does not select specific regions by PCR and detects microorganisms without hypotheses. Metagenomic analysis has mainly used next-generation sequencing (short-read sequencing), such as Miseq provided by Illumina. The Nanopore MinION™, a portable, mouse-sized, artificial intelligence (AI)-equipped long-read sequencer that can rapidly sequence whole genomes offered by ONT (Oxford Nanopore Technologies), has been attracting attention as a new sequencing platform.^13,14,^

Genomic analysis has the following advantages over culture methods: 1) The ability to obtain information regarding the species of the inflammable microorganisms and drug resistance in a short period enables the rapid introduction of effective treatment. 2) It can detect microorganisms difficult to culture and some that are not expected. 3) Detailed microorganism classification is possible. In 2017, Charalampous et al. compared the detection rate of culture, which is the gold standard for clinical diagnosis of bacterial lower respiratory tract infections, and genomic analysis, which combines metagenomic analysis using nanopores and qPCR, and reported that genomic analysis exhibited almost 100% sensitivity and specificity compared to the culture method.^12^ These facts indicate that metagenomic analysis can become the new standard for infectious disease diagnosis.

There are a few reports on the Nanopore application to ophthalmic infectious diseases,^5,6^ and no study have compared the sensitivity of nanopore long-read sequencing with mPCR till date. Therefore, we performed long read sequencing of samples obtained from the anterior aqueous humor of patients with infectious uveitis whose pathogens could be identified by mPCR and attempted to detect the etiologic microorganisms by metagenomic analysis.

## Methods

### Study design

This was a retrospective, observational case series study. We followed the tenets of the Declaration of Helsinki, received ethical approval from the Institutional Review Board of Nagoya University Graduate School of Medicine (2020-0598), and registered the study with the University Hospital Medical Information Network (UMIN000044906). Additionally, we obtained written informed consent from all patients for the testing of the samples. All participants were recruited at Nagoya University Hospital, Tokyo Medical University Hospital, or Yokkaichi Municipal Hospital, and patient clinical information was obtained retrospectively from medical records.

### Sample collection

This study included consecutive patients diagnosed with unilateral or bilateral anterior uveitis or panuveitis based on a comprehensive ophthalmological examination and clinical history. Only eyes that underwent mPCR testing to assess herpesvirus-related infections were analyzed. The remaining anterior aqueous humor samples were used for mPCR testing in patients with uveitis. Patients were excluded if they had other intraocular inflammation types, such as postprocedural endophthalmitis or noninfectious uveitis (e.g., sarcoidosis, Vogt-Koyanagi-Harada disease, or Behçet disease). Control subjects had cataracts, and the anterior aqueous humor samples were collected from patients with the cataracts at the beginning of the surgical procedure. All samples were collected and stored at −80°C between April 2017 and January 2023.

### DNA purification and nanopore long-read sequencing protocol

The QIAamp MinElute Virus Spin kit (Qiagen, Hilden, Germany) was used to purify and extract viral genomic DNA from anterior aqueous humor samples according to the manufacturer’s protocol. To procure a sufficient amount of DNA for analysis, a protocol using whole genome amplification was formulated (**Figure 1**). In summary, 10–20 µl of the collected anterior aqueous humor was subjected to DNA purification without isolation culture and whole genome amplification using the REPLI-g^®^ UltraFast kit. DNA quantification was performed using the high sensitivity dsDNA assay kit (Thermo Fisher-Q32851) on the Qubit 4.0 Fluorometer (Thermo Fisher-Q33238). DNA quality and fragment size (PCR products and MinION libraries) were assessed using the TapeStation 4150 (Agilent Technologies-G2992AA) automated electrophoresis platform with the Genomic ScreenTape (Agilent Technologies-5067-5365) and a DNA ladder (200 to >60,000 bp, Agilent Technologies-5067-5366). Long-read sequencing using the Flongle Flow Cell, Nanopore’s dedicated measurement chip, was then performed to evaluate the phylogenetic microorganism composition in the specimens. ONT MinKNOW software (v.22.05.5) was used to collect raw sequencing data. The EPI2MELabs Jupyter notebook server v1.1.6 (Oxford Nanopore Technologies) was used for the initial analysis of MinION data for the identification of pathogenic microorganisms present in the sample. Further downstream analysis for genome coverage was performed using minimap2 with default parameters for long-read data (-a -x map-ont). The minimum time to obtain analysis results using this method was approximately 190 min.

**Figure 1.**
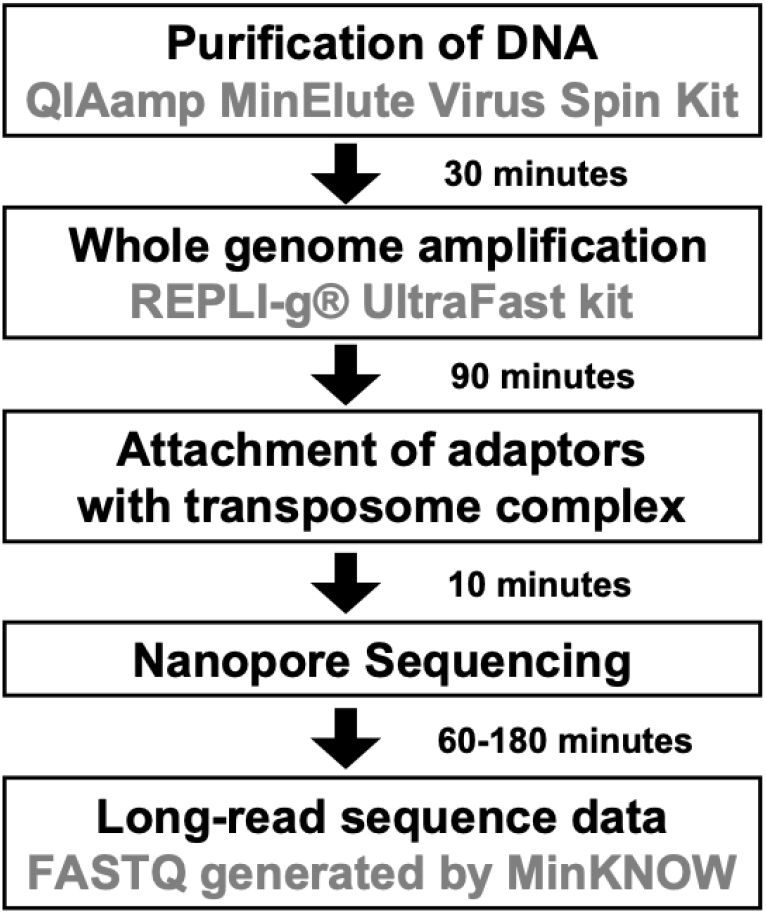
Nanopore long-read sequencing protocol for anterior aqueous humor samples for ocular infections. The anterior aqueous humor (10–20 µl) is subjected to whole genome amplification after DNA purification. The DNA is then treated with transposase and an adapter is added, and rapid long-read sequencing is performed using the Flongle Flow Cell, a nanopore-specific measurement chip, to evaluate the phylogenetic composition of the microorganisms in the specimen. Analysis results can be obtained in as little as 190 min after specimen collection using this method.

### Statistical analysis

The Fisher exact test was used for the categorical variables. We determined the significance of the differences in the copies of virus DNA between Nanopore positive and negative group by Kruskal-Wallis tests. We considered a *P* value of <0.05 statistically significant. Bonferroni-corrected *p*-values for multiple comparisons were applied. The R software version 3.4.4 was used for all statistical analyses.

## Results

### Phylogenetic classification and visualization of metagenomic analysis

A total of 22 anterior aqueous humor samples from uveitis patients with pathogens identified by mPCR were subjected to analysis (**Table 1, 2** and **eFigure 1**). The varicella zoster virus (VZV), cytomegalovirus (CMV), and Epstein-Barr virus (EBV) were detected using the metagenome analysis tool (EPI2ME) on nanopore sequence data, and the results of phylogenetic analysis were visualized (**Figure 2** and **eFigure 1**)^22^. The left side of the phylogenetic tree shows all microorganisms, except the human genome, and as one moves to the right, more detailed classifications such as domain (D: Domain), phylum (P: Phylum), class (C: Class), family (F: Family), genus (G: Genus), and species (S: Species) are quantitatively presented. The phylogenetic analyses of CMV retinitis (**Figure 2A**) are shown as representative examples. Reads of DNA fragments derived from the virus detected in each case were mapped to the reference genome of each virus.^15,17,18^ The DNA fragments of CMV were detected approximately 30 min after sequencing began in a case of CMV retinitis, and diagnosis was considered possible at this point (**Figure 2B**).

**Table 1.** Clinical characteristics of controls and patients with mPCR-positive, mPCR-negative uveitis.

|  | **mPCR-positive**<br>**uveitis (N = 22)** | **mPCR-negative**<br>**uveitis (N = 23)** | **Controls**<br>**(N = 22)** |
| --- | --- | --- | --- |
| **Age, mean (SD), y** | 60.4 (13.2) | 56.3 (14.8) | 71.7 (4.1) |
| **Gender, No. (%)** |  |  |  |
| **Male** | 11 (50.0) | 15 (65.2) | 12 (54.5) |
| **Female** | 11 (50.0) | 8 (34.8) | 10 (45.5) |
| **Types of herpes viruses**<br>**detected in the aqueous**<br>**humor, No. (%)** |  |  |  |
| **HSV-1** | 2 (9.1) | – | – |
| **HSV-2** | 1 (4.5) | – | – |
| **VZV** | 9 (40.9) | – | – |
| **EBV** | 1 (4.5) | – | – |
| **CMV** | 9 (41.0) | – | – |
| **Type of uveitis, No. (%)** |  |  |  |
| **Panuveitis** | 12 (54.5) | 12 (52.2) | – |
| **Anterior** | 10 (45.5) | 11 (47.8) | – |
| **Intermediate** | – | – | – |
| **Posterior** | – | – | – |

Abbreviations: CMV, cytomegalovirus; EB, Epstein-Barr virus; HSV, herpes simplex virus; mPCR, multiplex polymerase chain reaction; VZV, varicella-zoster virus.

**Table 2.** Result of the nanopore metagenomic analysis for mPCR-positive samples.

| ID | Gender | Age | Disease | Virus | DNA copy number | Nanopore metagenomic analysis |
| --- | --- | --- | --- | --- | --- | --- |
| P3 | F | 50's | ARN | VZV | unmeasured | Positive |
| P5 | M | 60's | CMV retinitis | CMV | unmeasured | Positive |
| P6 | F | 70's | CMV retinitis | CMV | unmeasured | Positive |
| P7 | F | 60's | CMV retinitis | CMV | unmeasured | N.D. |
| P8 | M | 60's | Herpetic iridocyclitis | CMV | unmeasured | N.D. |
| P9 | M | 60's | Herpetic iridocyclitis | CMV | unmeasured | N.D. |
| P10 | M | 50's | CMV retinitis | CMV | $9.6 \times 10^5$ | Positive |
| P11 | M | 70's | CMV retinitis | CMV | $5.8 \times 10^2$ | N.D. |
| P12 | F | 40's | CMV retinitis | CMV | unmeasured | N.D. |
| P13 | F | 60's | CMV retinitis | CMV | $4.2 \times 10^3$ | Positive |
| P18 | M | 70's | Herpetic iridocyclitis | HSV-1 | $7.1 \times 10^3$ | N.D. |
| P19 | F | 50's | Herpetic iridocyclitis | HSV-2 | $7.3 \times 10^3$ | N.D. |
| P20 | M | 60's | Herpetic iridocyclitis | VZV | $7.3 \times 10^5$ | Positive |
| P21 | M | 50's | ARN | VZV | $2.4 \times 10^5$ | N.D. |
| P22 | M | 60's | Herpetic iridocyclitis | VZV | $3.9 \times 10^8$ | Positive |
| P26 | F | 40's | Herpetic iridocyclitis | VZV | $1.1 \times 10^6$ | Positive |
| P27 | M | 20's | ARN | VZV | $4.1 \times 10^5$ | Positive |
| P29 | M | 40's | Herpetic iridocyclitis | VZV | $3.2 \times 10^5$ | Positive |
| P30 | F | 40's | Herpetic iridocyclitis | VZV | $3.5 \times 10^5$ | Positive |
| P31 | F | 80's | ARN | VZV | $3.9 \times 10^6$ | Positive |
| U14 | F | 60's | EBV malignant lymphoma | EBV | unmeasured | Positive |
| U242 | F | 80's | Herpetic iridocyclitis | HSV-1 | unmeasured | N.D. |

Abbreviations: mPCR, multiplex polymerase chain reaction; CMV, cytomegalovirus; EB, Epstein-Barr virus; HSV, herpes simplex virus; VZV, varicella-zoster virus: N.D., not detected.

**Figure 2.**
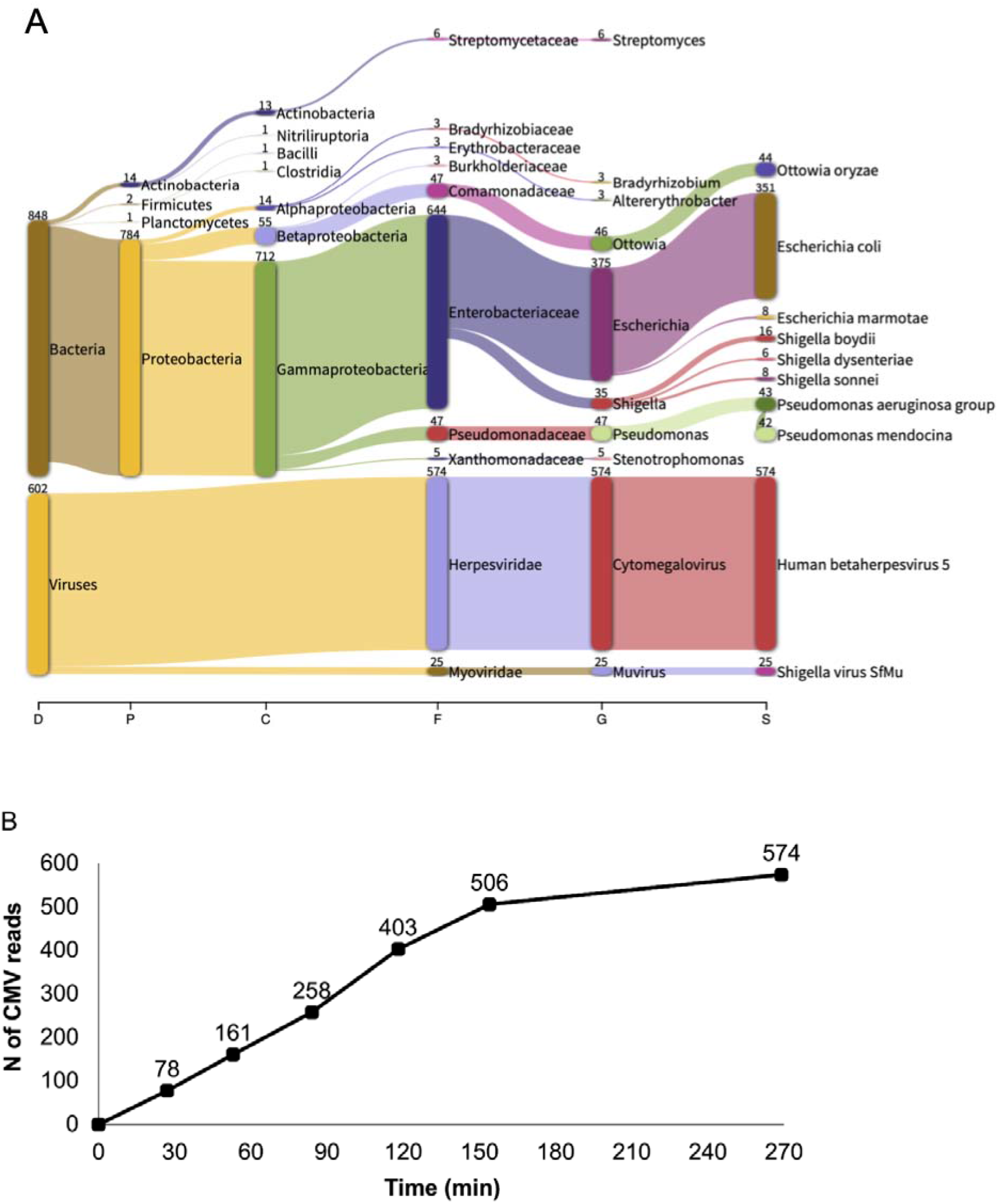
Phylogenetic classification of representative herpesvirus uveitis specimens and time-point analysis of the number of sequence reads. (A) Analysis results of case with CMV retinitis (P5), which is a representative of herpes virus uveitis. The phylogenetic tree (Pavian plot) shows all microorganisms except the human genome, and from left to right, domain (D: Domain), phylum (P: Phylum), class (C: Class), family (F: Family), genus (G: Genus) and species (S: Species) are quantified. (B) Time-point analysis of the number of sequence reads mapped to the CMV reference genome. The horizontal axis indicates time (minutes) and the vertical axis indicates the number of sequence reads mapped to the reference genome of CMV. CMV DNA reads were detected in about 30 min immediately after sequencing initiation.

### Limits of detection (LOD) of herpes viruses in nanopore sequencing

The detection rate of the etiologic virus for the 22 mPCR-positive samples in the nanopore metagenomic analysis was approximately 59.0% (**Table 2**). The cases that could not be detected tended to have significantly lower copy numbers by mPCR (P = 0.02), which is consistent with the lower metagenomic analysis sensitivity compared to mPCR. The LOD was between 10^3.6^ and 10^6^ copies of virus DNA (**Figure 3**). In the uveitis cases that were negative by mPCR (23 cases), the nine pathogenic microorganisms evaluated by mPCR were also not detected by nanopore in all cases.

**Figure 3.**
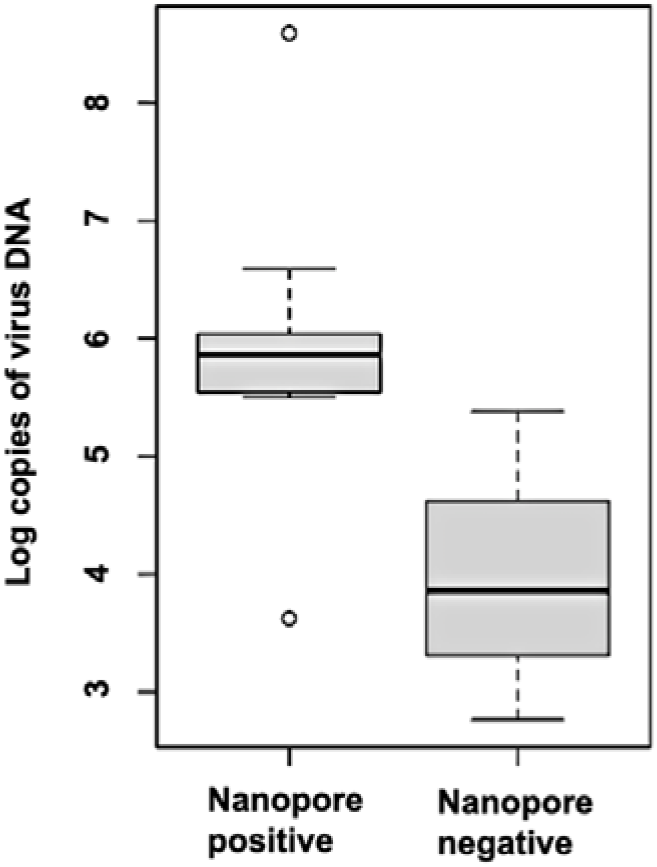
Limits of detection (LOD) of herpesviruses in nanopore sequencing. The number of copies of the target virus in mPCR-positive cases was compared among the Nanopore-positive and -negative groups. The number of copies in the Nanopore-positive group was significantly higher than that in the Nanopore-negative group (P = 0.02).

In contrast, nanopore also revealed that numerous bacteria and other microorganisms besides pathogens were detected in the phylogenetic analysis results (**Figure 2**). It is assumed that most of them are contaminants because several of the detected bacteria include *E. coli* and other bacteria known to cause severe infectious endophthalmitis. Many bacteria and other microorganisms that should not be present were confirmed and detected when sterile saline solution for medical use was treated in a sterile environment and analyzed by nanopore (**Figure 4**). This was reproducible even after multiple follow-ups and was considered to be due to reagent contamination from the environment and host.^8^ This suggests that our nanopore metagenome analysis results contain considerable noise, and that contamination control is necessary.

**Figure 4.**
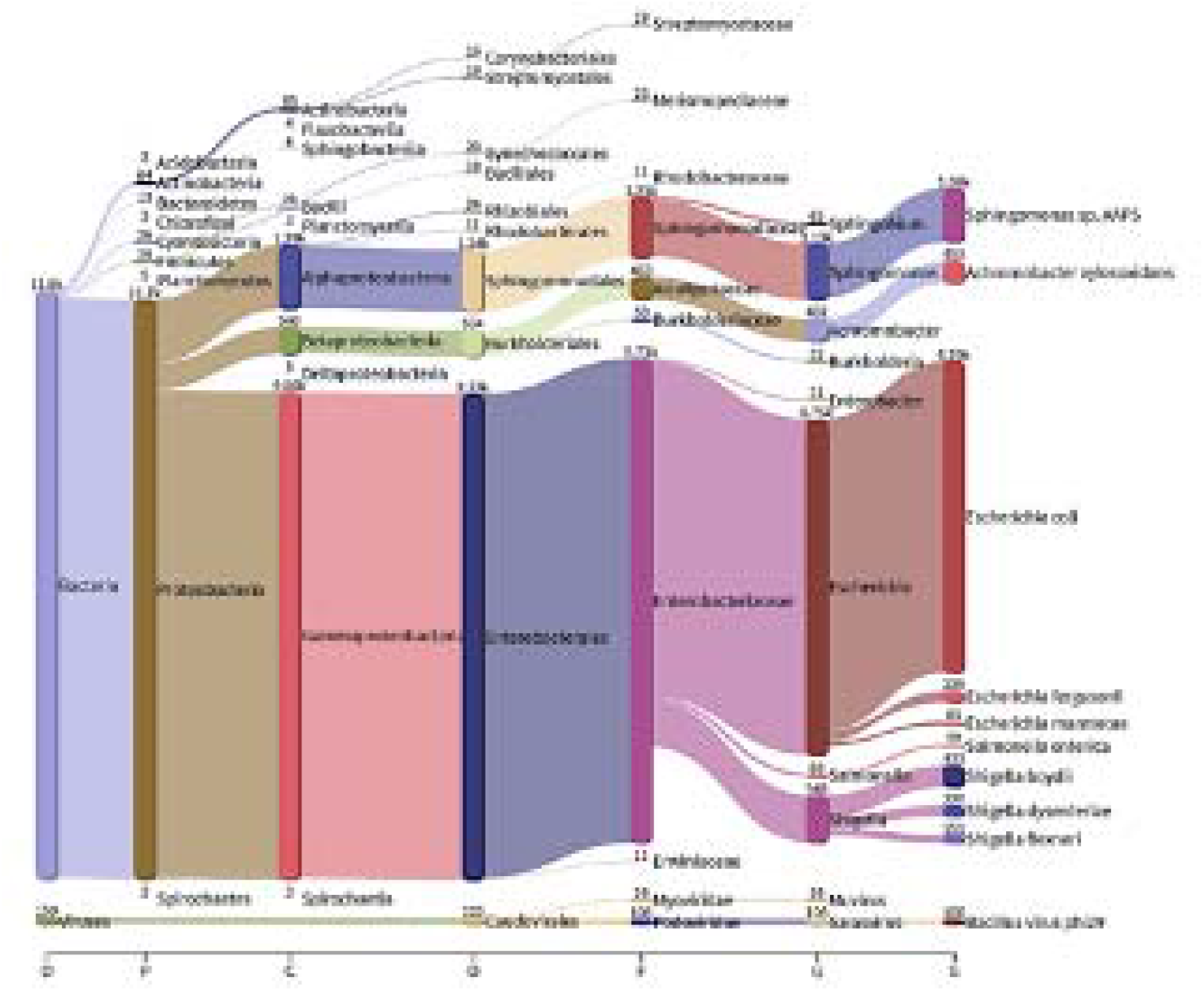
Sequencing results of sterile saline solution. A large number of microorganisms were detected when a sterile saline solution for medical use was treated in a sterile environment and analyzed by nanopore. This was considered to be due to contamination from reagents, the environment, and the host.

### Metagenomic association analysis as contamination control and pathogen discovery

To identify pathogens among the many contaminating microorganisms in the comprehensive analysis, we performed an additional metagenomic association analysis for contamination control and pathogen discovery. The results of nanopore metagenomic analysis were compared between groups of 23 cases of undiagnosed uveitis (mPCR-negative group) and 22 cases of cataract (cataract group). The same metagenomic analysis was performed on saline (n = 5) to remove contamination, and the detected microorganisms were interpreted as noise and used for data cleaning. An association analysis was performed between the mPCR-negative and cataract groups using the class-level classification in the detected phylogenetic tree to search for microorganisms commonly present in the former group. A total of 21 microorganisms were detected in both groups after noise was removed with saline data (**eTable 1**). Statistical group comparison between cataract and mPCR-negative cases showed that *Chlamydiae* was positive in 10/23 cases in the mPCR-negative group and 0/22 cases in the cataract group (P = 6.0 × 10^−4^) and was also significantly more frequent under multiple testing correction (significance level: *P* < 2.5 × 10^−3^ [=0.05/20]). No other microorganisms were observed to be detected significantly in either group. However, sequence reads from *Chlamydiae*-positive samples were biasedly mapped to a common region of the *Chlamydia trachomatis* subtype genome, which coincided with the *tet(C)* region of tetracycline (TC) resistance gene with low species specificity (**eFigure 2**). Furthermore, PCR was performed using specific and commonly available primers for *Chlamydia* which yielded negative results in mPCR-negative cases (data not shown).

## Discussion

In this study, we developed a sequencing protocol to detect the DNA fragments of etiologic viruses from the anterior aqueous humor of patients with herpes virus uveitis. Conversely, nanopore metagenomic results contained considerable noise and were found to be less sensitive compared with conventional mPCR tests for ocular infections, indicating that different characteristics between mPCR testing and nanopore metagenomic analysis (**Table 3**). mPCR requires a known pathogen targeting but has very high sensitivity and specificity. Nanopore metagenomic analysis allows for comprehensive analysis; however, it detects many microorganisms, including contaminants. Therefore, the interpretation of the results is difficult, and its sensitivity to known pathogens is clearly lower than that of mPCR.

**Table 3.** The strengths and weaknesses of Multiplex qPCR and nanopore sequencing.

|  | **Multiplex qPCR** | **Nanopore sequencing** |
| --- | --- | --- |
| **Strength** | High sensitivity & specificity | Comprehensive search for pathogen<br>Antibiotics resistance |
| **Weakness** | Predefined pathogen<br>Small information | Low sensitivity<br>Noisy data |

Because of these characteristics, it is necessary to identify pathogens among numerous contaminating microorganisms in the comprehensive analysis of pathogens. Therefore, we performed metagenomic association analysis for contamination control and pathogen discovery. Nanopore metagenomic association analysis detected the *tet(C)* region of the *Chlamydia trachomatis* genome was significantly more frequent in the former group. The *tet(C)* is a tetracycline efflux pump found in many species of Gram-negative bacteria, and the reported resistant strains is *Chlamydia trachomatis*^21^. However, this region had low species specificity in the blast search, and the genome was not amplified by PCR using additional *Chlamydia trachomatis*-specific primers.^16^ This suggests that the microorganism detected was not *Chlamydia trachomatis*, and the identity of the genome associated with mPCR-negative uveitis detected in the metagenomic association analysis has not been determined. A multifaceted genomic analysis, including whole genome sequencing is currently underway to identify the pathogen from which the detected genomic fragments were derived^19^.

Another issue in conducting comprehensive metagenomic analysis is the establishment of methods to remove human genomes. Approximately 99% of the acquired data is of the human genome, and if the human genome can be removed during sequencing, the amount of genomic information on pathogenic microorganisms to be detected will relatively increase.^9-11^ ONT is currently developing a system called adaptive sampling, which enriches and increase the desired sequence information by using an analysis program that automatically stops when unnecessary genomic information is detected and moves on to decipher another genome when sequencing with Nanopore MinION™ or other technologies. This method is used to enrich and increase the desired sequence information. Using this method, we increased the number of microbial genome reads by drastically reducing the number of unnecessary human genome analyses and thereby increasing the sensitivity of pathogenic microorganism detection and optimizing metagenome interpretation^20^.

In conclusion, our established sequencing protocol from the anterior aqueous humor detected the DNA fragments of etiologic viruses of typical herpes virus uveitis. However, the nanopore metagenomic results contained considerable noise and were found to be less sensitive compared to the conventional mPCR tests for ocular infections. Further study is needed to increase the amount of data on microbial genomes by removing human genomes and establishing contamination control in comprehensive metagenomic analysis.

## Supporting information

eFigure1

eFigure2

## Data Availability

All data produced in the present study are available upon reasonable request to the authors.

## Acknowledgments

Yoshito Koyanagi and Ai Fujita Sajiki contributed equally as co–first authors. Yoshito Koyanagi and Hiroaki Ushida are co–corresponding authors. Yoshito Koyanagi is the primary corresponding author and is responsible for sample analysis. Hiroaki Ushida is responsible for clinical information. This study was supported by a grant from the Japan Society for the Promotion of Science KAKENHI, grant number 20K09765 (Koji M Nishiguchi) and 22K20958 (Ai Fujita Sajiki).

## Figure legends

**eFigure 1. Summary of metagenomic profiling results for all the participants with mPCR-positive uveitis**

Summary of rapid metagenomic profiling results in all participants with mPCR-positive uveitis were shown. The phylogenetic tree (Pavian plot) shows all microorganisms except the human genome and from left to right, domain (D: Domain), phylum (P: Phylum), class (C: Class), family (F: Family), genus (G: Genus) and species (S: Species) are quantified.

**eFigure 2. *Tet(C)* region of the tetracycline (TC) resistance gene on the reference genome of a subtype of *Chlamydia trachomatis* mapped by Nanopore sequence reads**

Sequence reads from *Chlamydiae*-positive samples (P-26) were mapped to the genome of the *Chlamydia trachomatis* subtype and visualized in the Integrative Genomics Viewer. Sequence reads from *Chlamydiae*-positive samples are biasedly mapped to a common region of the genome. The region was searched with the Basic Local Alignment Search Tool and matched the *tet(C)* region of the TC gene, which has low species specificity.

## Tables

**eTable 1.**
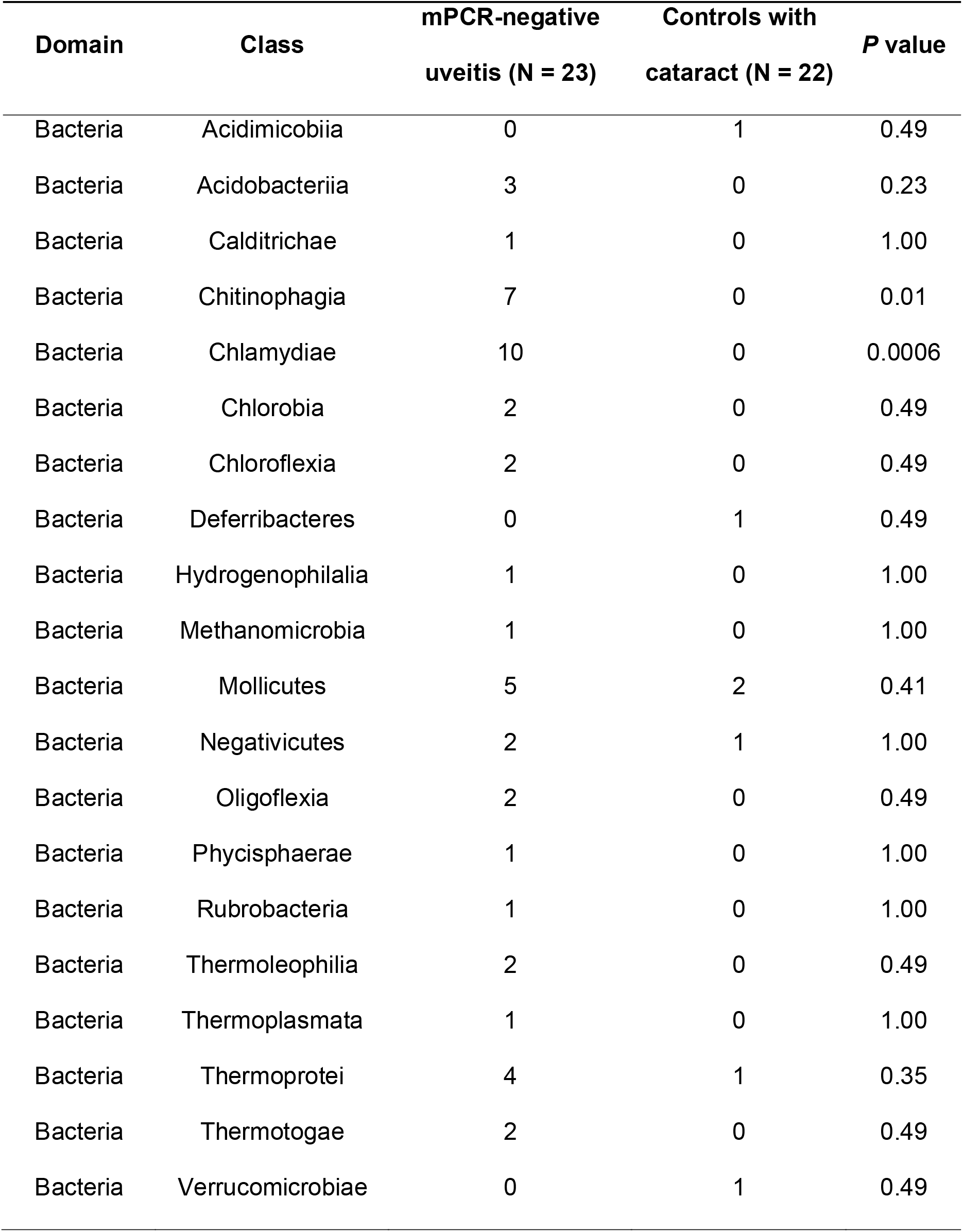
Result of metagenomic association analysis.

## Reference

1. Sugita S, Shimizu N, Watanabe K, Mizukami M, Morio T, Sugamoto Y, et al: Use of multiplex PCR and real-time PCR to detect human herpes virus genome in ocular fluids of patients with uveitis. Br J Ophthalmol 92: 928–932, 2008.

2. Sugita S, Ogawa M, Shimizu N, Morio T, Ohguro N, Nakai K, et al: Use of a comprehensive polymerase chain reaction system for diagnosis of ocular infectious diseases. Ophthalmology 120: 1761–1768, 2013.

3. Nakano S, Tomaru Y, Kubota T, Takase H, Mochizuki M, Shimizu N, et al: Multiplex Solid-Phase Real-Time Polymerase Chain Reaction without DNA Extraction: A Rapid Intraoperative Diagnosis Using Microvolumes. Ophthalmology 128: 729–739, 2021.

4. Sonoda KH, Hasegawa E, Namba K, Okada AA, Ohguro N, Goto H. Epidemiology of uveitis in Japan: a 2016 retrospective nationwide survey. Jpn J Ophthalmol. 65: 184–190. 2021.

5. Low L, Nakamichi K, Akileswaran L, Lee CS, Lee AY, Moussa G, et al. Deep Metagenomic Sequencing for Endophthalmitis Pathogen Detection Using a Nanopore Platform. Am J Ophthalmol. 242: 243–251. 2022.

6. Ishino M, Omi M, Araki-Sasaki K, Oba S, Yamada H, Matsuo Y, et al. Successful identification of Granulicatella adiacens in postoperative acute infectious endophthalmitis using a bacterial 16S ribosomal RNA gene-sequencing platform with MinION−: A case report. Am J Ophthalmol Case Rep. 26: 101524. 2022.

7. Salter SJ, Cox MJ, Turek EM, Calus ST, Cookson WO, Moffatt MF, et al. Reagent and laboratory contamination can critically impact sequence-based microbiome analyses. BMC Biol. 12: 87. 2014.

8. Marquet M, Zöllkau J, Pastuschek J, Viehweger A, Schleußner E, Makarewicz O, et al. Evaluation of microbiome enrichment and host DNA depletion in human vaginal samples using Oxford Nanopore’s adaptive sequencing. Sci Rep. 12: 4000. 2022.

9. Gan M, Wu B, Yan G, Li G, Sun L, Lu G, et al. Combined nanopore adaptive sequencing and enzyme-based host depletion efficiently enriched microbial sequences and identified missing respiratory pathogens. BMC Genomics. 22: 732. 2021.

10. Lin Y, Dai Y, Liu Y, Ren Z, Guo H, Li Z, et al. Rapid PCR-Based Nanopore Adaptive Sequencing Improves Sensitivity and Timeliness of Viral Clinical Detection and Genome Surveillance. Front Microbiol. 13: 929241. 2022.

11. Charalampous T, Kay GL, Richardson H, Aydin A, Baldan R, Jeanes C, et al. Nanopore metagenomics enables rapid clinical diagnosis of bacterial lower respiratory infection. Nat Biotechnol. 37: 783–792. 2019.

12. Kasianowicz JJ, Brandin E, Branton D, Deamer DW. Characterization of individual polynucleotide molecules using a membrane channel. Proc Natl Acad Sci U S A. 93: 13770–3. 1996.

13. Howorka S, Cheley S, Bayley H. Sequence-specific detection of individual DNA strands using engineered nanopores. Nat Biotechnol. 19: 636–9. 2001.

14. Robinson JT, Thorvaldsdóttir H, Winckler W, Guttman M, Lander ES, Getz G, Mesirov JP. Integrative genomics viewer. Nat Biotechnol. 29: 24–6. 2011.

15. Altschul SF, Gish W, Miller W, Myers EW, Lipman DJ. Basic local alignment search tool. J Mol Biol. 215: 403–10. 1990.

16. O’Leary NA, Wright MW, Brister JR, Ciufo S, Haddad D, McVeigh R, et al. Reference sequence (RefSeq) database at NCBI: current status, taxonomic expansion, and functional annotation. Nucleic Acids Res. 44: 733–45. 2016.

17. Schoch CL, Ciufo S, Domrachev M, Hotton CL, Kannan S, Khovanskaya R, et al. NCBI Taxonomy: a comprehensive update on curation, resources and tools. Database (Oxford). 2020: baaa062. 2020.

18. Harris SR, Clarke IN, Seth-Smith HM, Solomon AW, Cutcliffe LT, Marsh P, et al. Whole-genome analysis of diverse Chlamydia trachomatis strains identifies phylogenetic relationships masked by current clinical typing. Nat Genet. 44: 413–9. 2012.

19. Meyer F, Fritz A, Deng ZL, Koslicki D, Lesker TR, Gurevich A, et al. Critical Assessment of Metagenome Interpretation: the second round of challenges. Nat Methods. 19: 429–440. 2022.

20. Benamri I, Azzouzi M, Sanak K, Moussa A, Radouani F. An overview of genes and mutations associated with Chlamydiae species’ resistance to antibiotics. Ann Clin Microbiol Antimicrob. 20: 59. 2021.

21. Breitwieser FP, Salzberg SL. Pavian: interactive analysis of metagenomics data for microbiome studies and pathogen identification. Bioinformatics. 36: 1303–1304. 2020.

