## Supplementary material for "Metagenomic profiling of long-read sequencing for clinical diagnosis of ocular inflammation": eFigure1

**eFigure 1. Summary of metagenomic profiling result in all participants with mPCR-positive uveitis.**

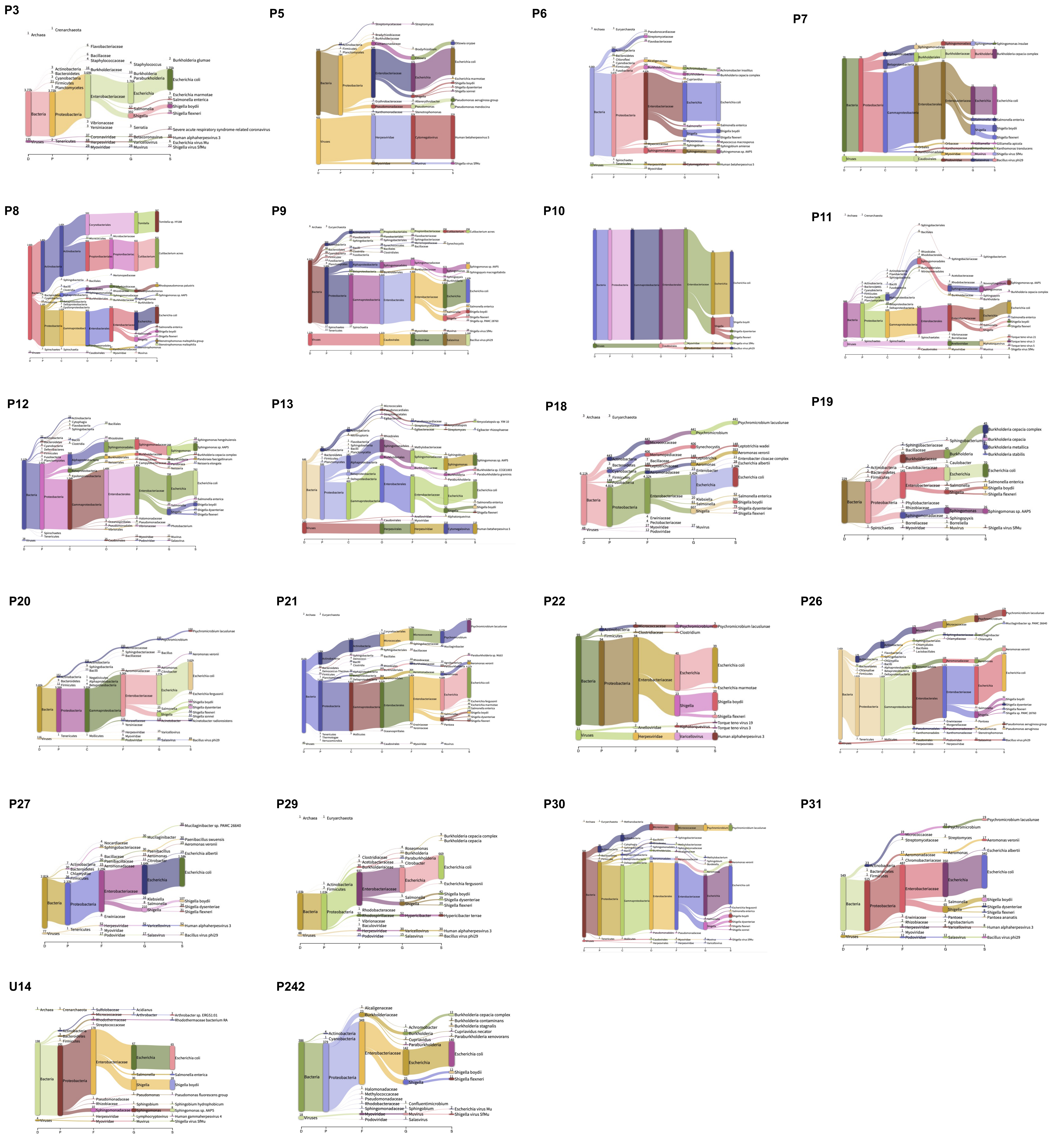


**Summary of metagenomic profiling result in all participants with mPCR-positive uveitis were shown. The phylogenetic tree (Pavian plot) shows all microorganisms except the human genome, and from left to right, domain (D: Domain), phylum (P: Phylum), class (C: Class), family (F: Family), genus (G: Genus) and species (S: Species) are quantified.**
