## Supplementary material for "Metagenomic profiling of long-read sequencing for clinical diagnosis of ocular inflammation": eFigure2

eFigure 2. *Tet(C)* region of the tetracycline (TC) resistance gene on the reference genome of a subtype of *Chlamydia trachomatis* mapped by Nanopore sequence reads.

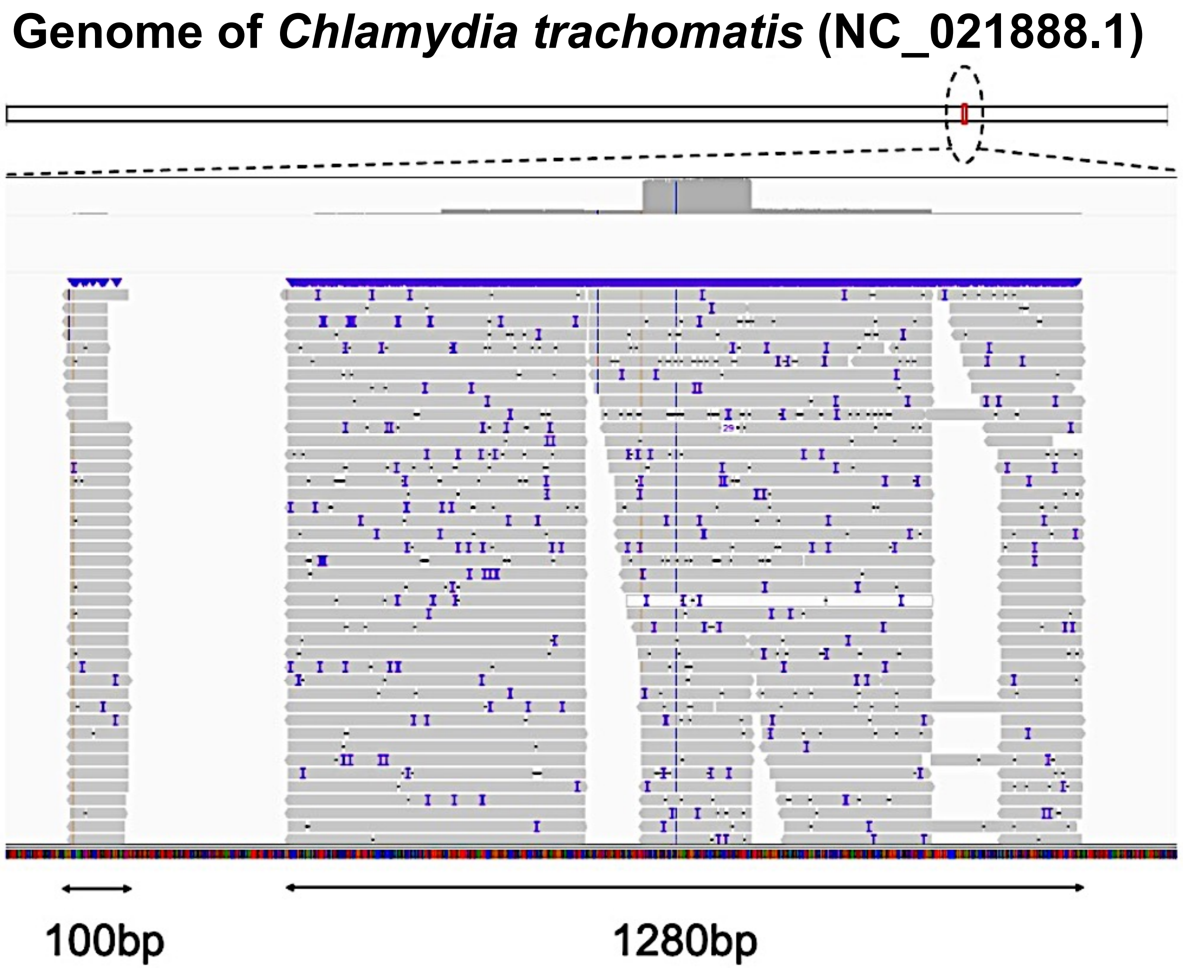


Sequence reads from *Chlamydiae-positive* samples (P-26) were mapped to the genome of the Chlamydia trachomatis subtype and visualized in the Integrative Genomics Viewer. Sequence reads from *Chlamydiae*-positive samples are biasedly mapped to a common region of the genome. The region was searched with the Basic Local Alignment Search Tool and matched the *tet(C)* region of the TC gene, which has low species specificity.
